# The impact of communicative and critical health literacy on trust in physicians among patients with systemic lupus erythematosus: the TRUMP2-SLE project

**DOI:** 10.1101/2022.05.13.22275070

**Authors:** Nao Oguro, Nobuyuki Yajima, Yoshia Miyawaki, Ryusuke Yoshimi, Yasuhiro Shimojima, Ken-ei Sada, Keigo Hayashi, Kenta Shidahara, Natsuki Sakurai, Chiharu Hidekawa, Dai Kishida, Takanori Ichikawa, Yuichi Ishikawa, Noriaki Kurita

## Abstract

**Objectives:** Accessing the Internet has increased the gap in patient health literacy (HL), impacting patient-doctor trust. We examined how trust in physicians is affected by functional HL (the ability to read and write) and by broader concepts of HL, including communicative HL (the ability to extract information from communication to use) and critical HL (the ability to analyze and use information) among patients with systemic lupus erythematosus (SLE).

**Methods:** This cross-sectional study enrolled 362 SLE patients at five academic centers between June 2020 and August 2021. The 14-item Functional Communicative Critical Health Literacy Scale assessed the three dimensions of HL (range: 1-4 points). Outcomes were trust in one’s physician and physicians generally using the 5-item Wake Forest Physician Trust Scale (range: 0-100 points). General linear models adjusted for age, sex, education, income, disease activity, disease duration, depression, and time using the Internet.

**Results:** Trust in one’s physician increased with higher functional and communicative HL (per 1-pt increase, 3.21 [95%CI 0.61, 5.81], 5.8 [95%CI 1.96, 9.63]). Trust in physicians in general increased with higher communicative HL and decreased with higher critical HL (per 1-pt increase, 7.01 [95%CI 2.27, 11.76], -6.83 [95%CI -11.67, -1.99]). Longer Internet use was associated with both higher communicative and critical HL.

**Conclusions:** Our findings suggest that rheumatologists can help patients build trust by encouraging dialogue about their health issues with their doctors and family members, rather than trying to improve their ability to discern health information.

**Key messages:**

- **What is already known on this topic** Factors affecting trust in one’s physician among chronic diseases including SLE are economic status, misdiagnosis experience, and the duration of the physician-patient relationship. In the general population, higher trust in physicians generally was associated with functional HL as patients’ external factors.
- **What this study adds** This is the first study to reveal an association between trust in one’s own physician and HL among patients with SLE. In addition, we were able to expand the correlates of HL by measuring broader concepts of HL dimensions, such as communicative and critical HL.
- **How this study may affect research, practice, or policy** This study suggests the importance of rheumatologists’ encouragement of patients to communicate, share their concerns, and resolve their misunderstandings to develop a trusting relationship. This study recommends that communictive HL (ability to obtain useful health information) should be the focus and not critical HL (ability to discern health information).

## INTRODUCTION

Trust in one’s physician among patients with systemic lupus erythematosus (SLE) constitutes the central component of the physician-patient encounter. [1, 2] Furthermore, it is essential for ensuring honest communication and confidence in decision making for continuation or change of life-long medication regimens or life events, such as birth control. Trust is not only based on competence but also on compassion for the patient and confidentiality, [1]and, therefore, is the key to facilitating the disclosure of a patient’s values and goals for care. Few studies have examined the factors affecting trust in one’s physician for chronic diseases, including SLE. Patient factors, such as economic status and misdiagnosis experience, [3] as well as relational factors, such as the duration of the physician-patient relationship, [3] are reported to be related to trust in one’s physician. However, whether disease-specific factors, such as disease activity, are reported to be associated with trust in physicians in general [2] and with trust in one’s physician, has not been studied. In addition, how external factors, such as exposure to information sources and health literacy (HL), affect trust in one’s physician among patients with SLE, has not been fully examined.

HL is narrowly viewed as the ability to sufficiently apply basic skills, such as reading and writing, to health-related information and is referred to as functional literacy. [4] A survey of the general population showed that higher functional HL was associated with greater trust in physicians. [5] However, trust in one’s physician was not examined and broader and more advanced skills of HL were not assessed. Much attention has also been paid to communicative literacy—the ability to extract information and meaning from several communications and to apply information to changing situations— and critical literacy—the ability to critically analyze information and use it to gain better control over life events and situations. [4, 6] SLE patients have frequent access to the Internet and blogs [7] and have lower trust in their physicians [8]. Thus, investigating how advanced HL and exposure to information resources among SLE patients affects trust in their physicians is valuable in building trust and creating dialogue to effectively navigate patients in choosing and maintaining appropriate treatment regimens.

Therefore, we aimed to examine how a variety of HL factors influence trust in one’s physician and trust in physicians, in general, using data from the TRUMP2-SLE (Trust Measurement for Physicians and Patients with SLE) study of Japanese SLE patients, in which patients and physicians were ethnically matched.

## METHODS

### Study design and setting

This was a cross-sectional study using baseline data from the TRUMP2-SLE study, an ongoing multicenter cohort study conducted at five academic medical centers (Showa University Hospital, Okayama University Hospital, Shinshu University Hospital, Yokohama City University Hospital, and Yokohama City University Medical Center). This study followed the Declaration of Helsinki and Good Clinical Practice guidelines and was approved by the Ethics Review Board of Showa University (number 22-002-A).

The inclusion criteria were as follows: (1) SLE patients aged 20 years or older, according to the revised 1997 American College of Rheumatology classification criteria, (2) receiving rheumatology care at the participating center, and (3) ability to respond to the questionnaire survey. Patients with dementia or total blindness were excluded from the study.

### Exposures

HL skills were measured using the original Japanese version of the Functional Communicative Critical Health Literacy Scale (FCCHL) by Ishikawa. [6] The FCCHL is a multidimensional construct that includes 14 items scored on a 4-point Likert scale. The FCCHL captures three domains: functional HL (five items), the ability to read or understand the instructions or leaflets from healthcare providers, hospitals, and pharmacies; communicative HL (five items), the ability to extract and communicate health information with doctors or the family; and critical HL (four items), the ability to critically analyze health information and use it to make decisions (Supplementary Table <u>HL and Trust in SLE - 6 Supplementary Table FCCHL.docx</u>). Patients were asked to score each item on a scale of 1 to 4, with 1 and 4 meaning “not at all” and “often,” respectively. Each domain was calculated as a mean score ranging from 1 (low HL) to 4 (high HL). The FCCHL was validated and demonstrated to have good reliability (coefficient alphas of 0.84, 0.77, and 0.65 for functional, communicative, and critical HL, respectively) and construct validity. [6]

### Outcomes

The main outcomes were “trust in one’s own physician” and “trust in physician’s generally”, which were measured by the 5-item Japanese version of the Wake Forest Physician Trust Scales (i.e., Interpersonal Trust in Physician Scale,’ and ‘Trust in Doctors Generally Scale,’ respectively. [9, 10]

Each scale was composed of five items scored using a 5-point Likert scale. Patients were asked to choose one of the following responses for each item: “strongly disagree” (1 point) to “strongly agree” (5 points). After inverting the score for a negatively worded item, the sum of the scores was converted into a scale ranging from 0 to 100.

The coefficient alpha for the Japanese version of the Interpersonal Trust in Physician Scale and the Trust in Doctors Generally Scale was 0.85 and 0.88, respectively, and demonstrated construct validity. [9]

### Measurement of covariates

Confounding variables were those suspected to determine both HL and trust in physicians, based on evidence in the literature. The variables included were age, sex, final education, household income, disease activity, duration of illness, depression, and time spent on the Internet. Disease activity was measured by the attending physician using the Systemic Lupus Erythematosus Disease Activity Index 2000 (SLEDAI-2K). Depression was measured by the single item “During the past 4 weeks, how often did you feel because of your lupus that you were depressed?” in the Japanese version of Lupus PRO and defined as the presence of depression when “None of the time” was not chosen. [11] Time spent on the Internet was measured by asking “How much time do you spend on the Internet and social network services in a day, not including time for work?” where respondents were asked to choose from six choices ranging from “never” to more than “four hours.” In the analysis step, the responses were merged into the following categories based on the distribution of the responses: “not at all,” “less than one hour,” “more than one hour to less than two hours,” and “more than two hours.” The questionnaire was administered at each facility between June 2020 and August 2021, and the patients were asked to complete it, either in the waiting room or at home. The questionnaire included assurances that the attending physician would not view the responses and that the responses would only be used at the central facility for aggregation.

### Statistical analysis

All statistical analyses were performed using Stata/SE, version 16.1 (StataCorp, College Station, TX, USA). Patient characteristics were described as frequencies and proportions for categorical variables and medians and interquartile ranges (IQR) for continuous variables. Histograms of trust in one’s physician and trust in physicians in general scores were constructed.

The associations between the abovementioned eight patient characteristics and each of the three domains of the FCCHL were analyzed using general linear models.

Next, the associations between the FCCHL and trust in one’s physician and trust in physicians in general were analyzed using general linear models. The eight patient characteristics described above were included in the multivariate analyses as covariates. Missing covariates were addressed using a multiple completion approach. Twenty imputations were performed by multiple imputations with chained equations, assuming that the analyzed data were missing at random. Statistical significance was set at p < 0.05.

### Patient and public involvement

Neither the general public nor patients with SLE were involved in the planning, recruitment and conduct of this study.

## RESULTS

### Study flow

Initially, 386 SLE patients who met the inclusion criteria were identified. Of these, 24 patients without an FCCHL score, trust in one’s physician score, or trust in physicians generally score were excluded. Overall, 362 patients were included in the analysis (Figure 1).

**Figure 1.**
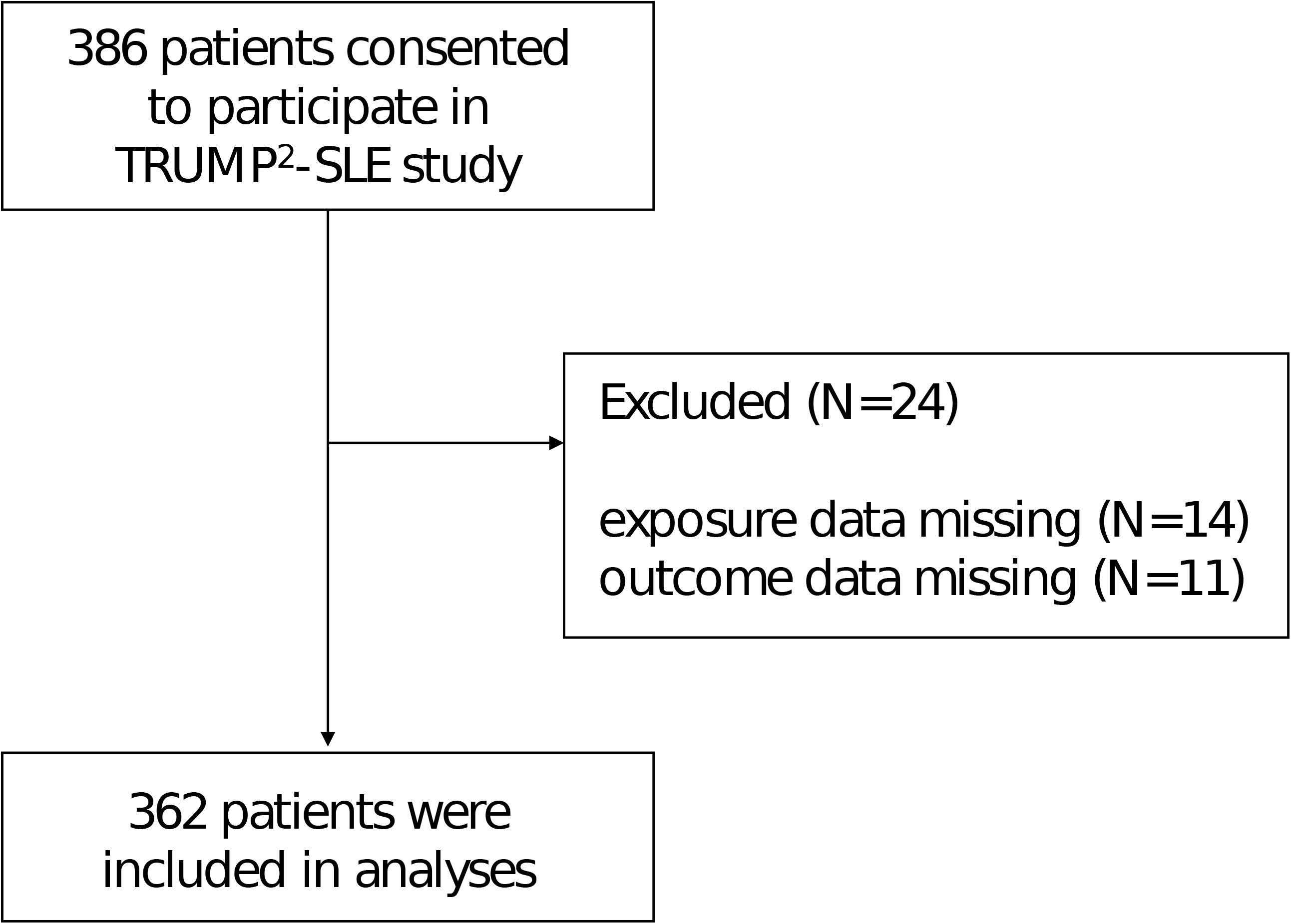
Flow diagram.

### Patient Characteristics

Patient characteristics in the primary analysis are presented in Table 1. The median age was 45 years (IQR 35-55) years and 319 (88%) were women. The median disease activity as determined by the SLEDAI-2K scale was 4.0 (IQR 2.0–8.0) points. Among HL domains (exposures), functional HL scores tended to be higher, followed by communicative HL and critical HL: 3.5 (IQR 3.0–4.0), 3.0 (IQR 2.5–3.4), and 2.8 (IQR 2.0–3.3). The median score of trust in one’s physician (outcome) was higher than that of general (outcome) physicians: 80 (IQR70–95) and 65 (IQR 50–80).

**Table 1.** Patients’ characteristics (n = 362)

|  | **Total**<br>**n = 362** |
| --- | --- |
| **<u>Demographics</u>** |  |
| Age, yr | 44.3 [35.3 - 54.1] |
| Women, n (%) | 319 (88.1 %) |
| **Education, n (%)** |  |
| Junior high school or lower | 17 (5 %) |
| High school/College | 243 (71.5 %) |
| University/Graduate school | 80 (23.5 %) |
| missing, n | 22 |
| **Household income, n (%)** |  |
| <1,000,000 yen | 27 (9.1 %) |
| 1,000,000 to <5,000,000 yen | 133 (44.6 %) |
| 5,000,000 to <10,000,000 yen | 118 (39.6 %) |
| >10,000,000 yen | 20 (6.7 %) |
| missing, n | 64 |
| **Disease duration, n (%)** |  |
| <5 yr, n (%) | 63 (18.1 %) |
| 5 yr to <10 yr, n (%) | 74 (21.2 %) |
| 10 yr to <20 yr, n (%) | 125 (35.8 %) |
| >20 yr, n (%) | 87 (24.9 %) |
| missing, n | 13 |
| **SLEDAI-2K, point** | 4 [2 - 8] |
| **Depressive symptom, n (%)** | 53 (18.1 %) |
| missing, n | 69 |
| **Duration of internet use, n (%)** |  |
| None | 49 (13.6 %) |
| <1 h | 124 (34.4 %) |
| 1 h to <2 h | 88 (24.4 %) |
| >2 h | 100 (27.7 %) |
| missing, n | 1 |
| **Health Literacy** |  |
| Functional HL | 3.5 [3 - 4] |
| Communicative HL | 3 [2.6 - 3.4] |
| Critical HL | 2.8 [2.3 - 3] |
| **Trust in Physician** |  |
| Trust in one's physician | 80 [70 - 95] |
| Trust in physicians in general | 65 [50 - 80] |

Continuous variables are summarized as median and interquartile range (in square brackets) (in parentheses). Categorical variables are summarized as frequencies and proportions (in parentheses). SLEDAI-2K: Systemic Lupus Erythematosus Disease Activity Index.

### Patient characteristics associated with health literacy

Table 2 shows the association between each domain of the FCCHL and patient characteristics. Functional HL was inversely associated with older age (per 10 yr increase, -0.1 [95%CI -0.17, -0.03]) and depressive symptoms (−0.31 [95%CI -0.52, -0.1]). Functional HL was positively associated with higher education levels (junior high school or lower and high school/college, 0.45 [95%CI 0.11, 0.79] and university/graduate school (0.5 [95%CI 0.14, 0.86]) and disease duration (less than 5 years and more than 5 years but less than 10 years, 0.24 [95%CI 0.02, 047]). Both communicative and critical HL were positively associated with longer Internet use (communicative HL vs. none: <1h, 0.37 [95%CI 0.13 to 0.61]; 1h to <2h, 0.56 [95%CI 0.3, 0.82]; >2h, 0.64 [95%CI 0.36, 0.91]; critical HL vs. none: <1h, 0.33 [95%CI 0.09, 0.57]; 1h to <2h, 0.6 [95%CI 0.35, 0.86]; >2h, 0.64 [95%CI 0.37, 0.91]).

**Table 2.** Associations of health literacy domains with covariates† (n = 362)

|  | Functional HL | Communicative HL | Critical HL |
| --- | --- | --- | --- |
|  | mean difference, point estimate<br>(95%CI) | mean difference, point estimate<br>(95%CI) | mean difference, point estimate<br>(95%CI) |
| **Age**, per 10-yr increase | **-0.1 (-0.17 to -0.03), p = 0.003** | 0.04 (-0.02 to 0.11), p = 0.17 | 0.02 (-0.05 to 0.08), p = 0.622 |
| Women vs. Men | 0.06 (-0.16 to 0.28), p = 0.571 | 0.1 (-0.11 to 0.31), p = 0.345 | -0.03 (-0.24 to 0.18), p = 0.783 |
| **Educational attainment** |  |  |  |
| Junior high school or lower | Reference | Reference | Reference |
| High school / College | **0.45 (0.11 to 0.79), p = 0.01** | -0.06 (-0.37 to 0.25), p = 0.719 | 0.03 (-0.28 to 0.33), p = 0.87 |
| University /<br>Graduate school | **0.5 (0.14 to 0.86), p = 0.006** | -0.16 (-0.49 to 0.18), p = 0.362 | -0.07 (-0.4 to 0.26), p = 0.676 |
| **Household income** |  |  |  |
| <1,000,000 yen | Reference | Reference | Reference |
| 1,000,000 to <5,000,000 yen | -0.01 (-0.28 to 0.25), p = 0.93 | -0.1 (-0.37 to 0.17), p = 0.447 | -0.05 (-0.33 to 0.23), p = 0.74 |
| 5,000,000 to <10,000,000 yen | 0.05 (-0.22 to 0.33), p = 0.701 | 0.06 (-0.22 to 0.34), p = 0.664 | 0.01 (-0.27 to 0.29), p = 0.934 |
| >10,000,000 yen | 0.18 (-0.21 to 0.57), p = 0.361 | 0.06 (-0.34 to 0.45), p = 0.773 | 0.04 (-0.36 to 0.43), p = 0.845 |
| **Disease duration** |  |  |  |
| <5 yr | Reference | Reference | Reference |
| 5 yr to <10 yr | **0.24 (0.02 to 0.47), p = 0.035** | 0.14 (-0.07 to 0.36), p = 0.196 | 0.12 (-0.09 to 0.33), p = 0.259 |
| 10 yr to <20 yr | 0.15 (-0.05 to 0.35), p = 0.152 | 0.14 (-0.06 to 0.33), p = 0.177 | 0.16 (-0.03 to 0.35), p = 0.098 |
| >20 yr | 0.03 (-0.2 to 0.26), p = 0.769 | 0.03 (-0.19 to 0.25), p = 0.762 | 0.1 (-0.11 to 0.32), p = 0.34 |
| **SLEDAI-2K**, per 1-pt increase | -0.01 (-0.02 to 0.01), p = 0.495 | 0 (-0.01 to 0.01), p = 0.955 | 0 (-0.01 to 0.02), p = 0.702 |
| **Depressive symptom** | **-0.31 (-0.52 to -0.1), p = 0.004** | 0.01 (-0.19 to 0.2), p = 0.954 | 0.06 (-0.15 to 0.26), p = 0.58 |
| **Duration of internet use** |  |  |  |
| None | Reference | Reference | Reference |
| <1 h | -0.02 (-0.26 to 0.23), p = 0.882 | **0.37 (0.13 to 0.61), p = 0.002** | **0.33 (0.09 to 0.57), p = 0.006** |
| 1 h to <2 h | -0.01 (-0.27 to 0.26), p = 0.963 | **0.56 (0.3 to 0.82), p <0.001** | **0.6 (0.35 to 0.86), p <0.001** |
| >2 yr | -0.02 (-0.31 to 0.26), p = 0.871 | **0.64 (0.36 to 0.91), p <0.001** | **0.64 (0.37 to 0.91), p <0.001** |

†For each health literacy domain score, general linear models were fitted with the inclusion of all the variables listed above.
SLEDAI-2K: Systemic Lupus Erythematosus Disease Activity Index.

### The association between health literacy and trust in physicians

Table 3 shows the association between HL status and trust in physicians. Trust in one’s physician increased with higher functional and communicative HL (per 1-pt increase: 3.21 [95%CI 0.61, 5.81] and 5.8 [95%CI 1.96, 9.63], respectively). Disease activity, measured as SLEDAI-2K, was also positively associated with higher trust in one’s physician (per 1-pt increase, 0.52 [95%CI 0.17, 0.88]). We found insufficient evidence that longer Internet use was associated with less trust in physicians.

**Table 3.** Associations of trust in patients’ rheumatologists with health literacy and covariates† (n = 362)

|  | Corresponding<br>standardized ES | mean difference, point<br>estimate (95%CI) | p-value |
| --- | --- | --- | --- |
| Functional HL, per 1-pt increase | **0.2** | **3.21 (0.61 - 5.81)** | **0.016** |
| Communicative HL, per 1-pt increase | **0.35** | **5.8 (1.96 - 9.63)** | **0.003** |
| Critical HL, per 1-pt increase | -0.09 | -1.48 (-5.38 - 2.42) | 0.455 |
| Age, per 10-yr increase | 0.03 | 0.41 (-1.21 - 2.03) | 0.618 |
| Women vs. Men | -0.3 | -4.92 (-10.23 - 0.39) | 0.069 |
| **Educational attainment** |  |  |  |
| Junior high school or lower |  | Reference |  |
| High school/College | -0.28 | -4.52 (-12.6 - 3.56) | 0.271 |
| University/Graduate school | -0.14 | -2.37 (-11.1 - 6.36) | 0.593 |
| **Household income** |  |  |  |
| <1,000,000 yen |  | Reference |  |
| 1,000,000 to <5,000,000 yen | -0.24 | -4 (-10.7 - 2.72) | 0.242 |
| 5,000,000 to <10,000,000 yen | -0.22 | -3.56 (-10.5 - 3.4) | 0.314 |
| >10,000,000 yen | -0.15 | -2.45 (-11.9 - 6.95) | 0.608 |
| **Disease duration** |  |  |  |
| <5 yr |  | Reference |  |
| 5 yr to <10 yr | 0.21 | 3.52 (-1.95 - 8.99) | 0.207 |
| 10 yr to <20 yr | 0.08 | 1.33 (-3.64 - 6.31) | 0.599 |
| >20 yr | -0.07 | -1.13 (-6.7 - 4.47) | 0.693 |
| **SLEDAI-2K, per 1-pt increase** | **0.03** | **0.52 (0.17 - 0.88)** | **0.004** |
| **Depressive symptom** | -0.05 | -0.81 (-5.72 - 4.1) | 0.746 |
| **Duration of internet use** |  |  |  |
| None |  | Reference |  |
| <1 h | 0.02 | 0.31 (-5.69 - 6.32) | 0.918 |
| 1 h to <2 h | -0.14 | -2.26 (-8.93 - 4.42) | 0.507 |
| >2 yr | -0.02 | -0.4 (-7.5 - 6.69) | 0.911 |

<sup>†</sup> The general linear model was fitted with the inclusion of all variables listed above. To calculate the corresponding standardized ES (Cohen's *d*), the point estimate was divided by the standard deviation of the trust in interpersonal physicians scores.
SLEDAI-2K: Systemic Lupus Erythematosus Disease Activity Index.

Table 4 shows the association between HL status and trust in physicians. Trust in doctors generally increased with higher communicative HL but decreased with higher critical HL (per 1-pt increase, 7.01 [95%CI 2.27, 11.76] and -6.83 [95%CI -11.67, -1.99], respectively). We found that women perceived less trust in physicians in general than did men (−8.7 [95%CI -15, -2.1]). Furthermore, longer Internet use was generally associated with less trust in physicians (vs. none: 1h to <2h, -9. 19[95%CI -17.5, -0.92] and >2h, -9.87 [95%CI -18.7, - 1.07]).

**Table 4.**
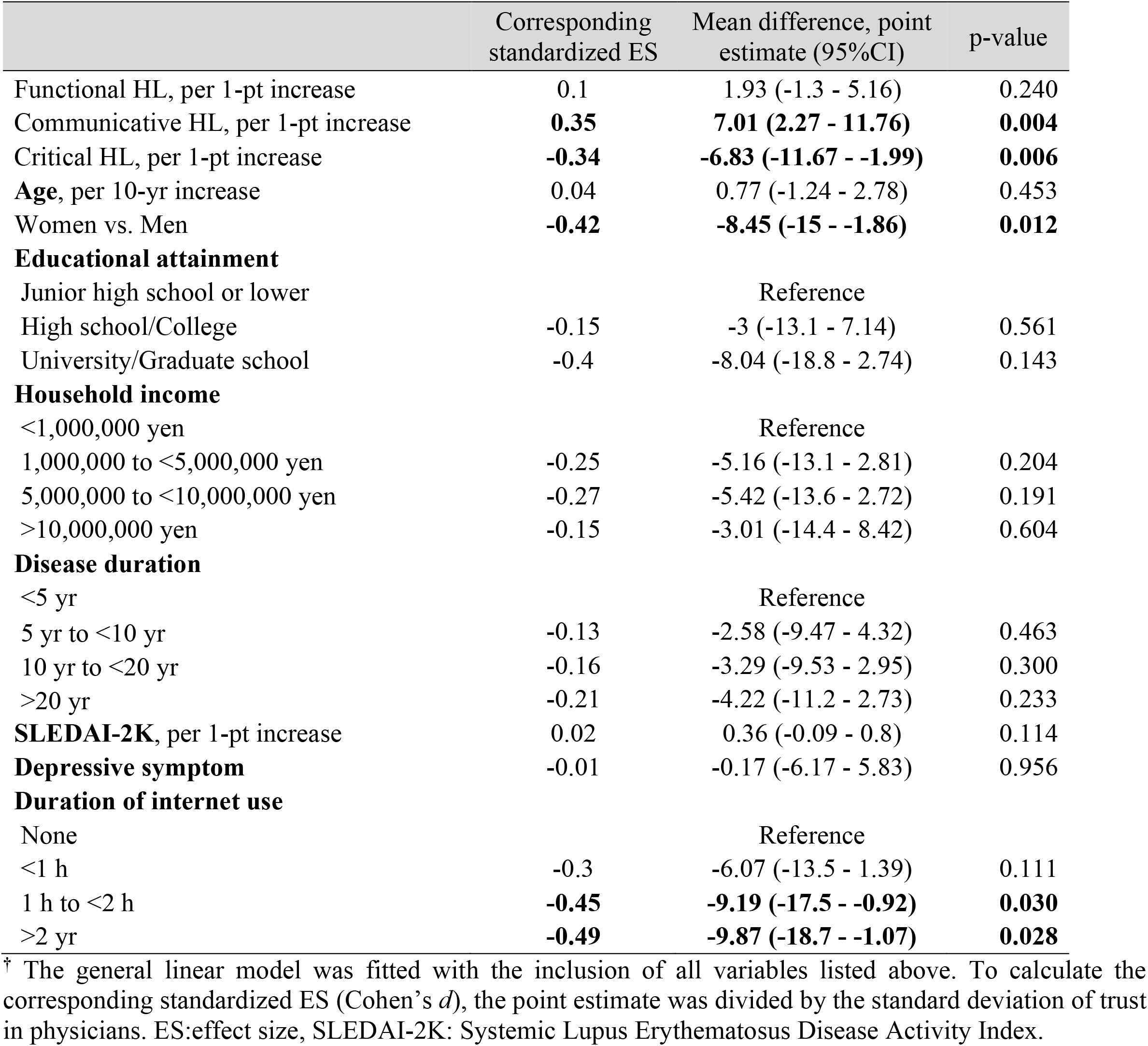
Associations of trust in general physicians with health literacy and covariates† (n = 362)

|  | Corresponding<br>standardized ES | Mean difference, point<br>estimate (95%CI) | p-value |
| --- | --- | --- | --- |
| Functional HL, per 1-pt increase | 0.1 | 1.93 (-1.3 - 5.16) | 0.240 |
| Communicative HL, per 1-pt increase | **0.35** | **7.01 (2.27 - 11.76)** | **0.004** |
| Critical HL, per 1-pt increase | **-0.34** | **-6.83 (-11.67 - -1.99)** | **0.006** |
| Age, per 10-yr increase | 0.04 | 0.77 (-1.24 - 2.78) | 0.453 |
| Women vs. Men | **-0.42** | **-8.45 (-15 - -1.86)** | **0.012** |
| **Educational attainment** |  |  |  |
| Junior high school or lower |  | Reference |  |
| High school/College | -0.15 | -3 (-13.1 - 7.14) | 0.561 |
| University/Graduate school | -0.4 | -8.04 (-18.8 - 2.74) | 0.143 |
| **Household income** |  |  |  |
| <1,000,000 yen |  | Reference |  |
| 1,000,000 to <5,000,000 yen | -0.25 | -5.16 (-13.1 - 2.81) | 0.204 |
| 5,000,000 to <10,000,000 yen | -0.27 | -5.42 (-13.6 - 2.72) | 0.191 |
| >10,000,000 yen | -0.15 | -3.01 (-14.4 - 8.42) | 0.604 |
| **Disease duration** |  |  |  |
| <5 yr |  | Reference |  |
| 5 yr to <10 yr | -0.13 | -2.58 (-9.47 - 4.32) | 0.463 |
| 10 yr to <20 yr | -0.16 | -3.29 (-9.53 - 2.95) | 0.300 |
| >20 yr | -0.21 | -4.22 (-11.2 - 2.73) | 0.233 |
| **SLEDAI-2K**, per 1-pt increase | 0.02 | 0.36 (-0.09 - 0.8) | 0.114 |
| **Depressive symptom** | -0.01 | -0.17 (-6.17 - 5.83) | 0.956 |
| **Duration of internet use** |  |  |  |
| None |  | Reference |  |
| <1 h | -0.3 | -6.07 (-13.5 - 1.39) | 0.111 |
| 1 h to <2 h | **-0.45** | **-9.19 (-17.5 - -0.92)** | **0.030** |
| >2 yr | **-0.49** | **-9.87 (-18.7 - -1.07)** | **0.028** |

<sup>†</sup> The general linear model was fitted with the inclusion of all variables listed above. To calculate the corresponding standardized ES (Cohen's *d*), the point estimate was divided by the standard deviation of trust in physicians. ES:effect size, SLEDAI-2K: Systemic Lupus Erythematosus Disease Activity Index.

## DISCUSSION

Our study aimed to clarify the association between the broader concept of HL (communicative and critical HL), which requires active attitudes to obtain, understand, analyze, and use health information, and trust in one’s physicians and physicians in general among SLE patients. Trust in physicians was significantly associated with high functional and communicative HL. Trust in physicians in general was associated with higher communicative HL but was inversely associated with critical HL.

Despite the recent increase in the importance of examining the impact of HL on the physician-patient relationship in patients with SLE, [12] our study is the first to analyze the associations in this specific population.

Specifically, our study showed that functional HL was associated with trust in physicians. In a survey of the general population in Taiwan, high functional HL was associated with higher trust in doctors. [5] In that study, trust in one’s physician was not examined; however, given that the study participants were from the general population, they did not necessarily have a disease and, therefore, did not have their own physician. Lack of functional HL in patients with SLE may lead to misunderstanding of health information and a loss of trust through conflicts between physicians’ recommendations for appropriate treatment and management and patients’ unrealistic expectations of incorrect management practices. [5] For example, there is a misunderstanding that as long as one takes corticosteroids at a lower dose than prescribed by a physician, the disease will not flare up. Acting on this belief may cause the disease to flare up which may compromise mutual trust if this patient’s behavior is not confirmed at the earliest clinical encounter.

Furthermore, the finding in our study that the association between higher communicative HL and higher trust in one’s physician was independent of educational level and functional HL is worth mentioning. This implies that communicative HL as a patient factor may influence trust in one’s physician through patient-physician dialogue, which cannot be explained by educational level or basic understanding. Given that higher communicative HL was also associated with higher levels of trust in doctors in general, it may reflect a basic ability to establish a trusting relationship with any physician.

The present findings reinforce the importance of modes of patient-physician dialogue in fostering trust. A previous study of patients with rheumatoid arthritis and SLE showed that a physician’s approach to patient-centered dialogue, such as considering the patient’s interests, was associated with higher trust in physicians in general. [2] This may indicate that communicative HL is also elicited by physicians through patient-centered dialogue, which leads patients to be more willing to disclose their concerns.

There are several explanations for the finding that higher critical HL was associated with lower trust in physicians in general but not with lower trust in one’s physician. First, patients with SLE may be more cautious about whether medical care provided by physicians other than their own rheumatologists is tailored specifically and appropriately to their disease.

For example, for contraception in the presence of antiphospholipid antibody syndrome, avoidance of pills may be necessary to minimize the risk of venous thrombosis. Moreover, for some regular medications, such as tacrolimus, avoidance of macrolide antibiotics may be necessary to avoid increased drug concentration due to drug interactions. Unless the attending rheumatologist is present, physicians may prescribe these drugs because they may not be aware that the patient has antiphospholipid antibody syndrome or is prescribed tacrolimus due to a lack of knowledge about them. In contrast, rheumatologists have a good understanding of patients’ individuality; therefore, a patient’s high critical HL may not necessarily lead to a decrease in trust in their rheumatologist.

Second, the finding in the present study that longer Internet use was associated with higher critical HL may reflect the reality that patients are increasingly accessing online health information, acquiring more knowledge about a variety of medical care, and comparing it to the actual medical care provided to them. The preference for obtaining online health information consistent with one’s beliefs among those with higher HL shown in a previous study may explain the decreased trust in physicians’ recommendations. [13]

This study has several clinical implications for rheumatologists and researchers. First, the development of methods to improve functional and communicative HL is warranted to enhance trust in physicians. Specifically, substituting the patient’s eyes and ears with help from a person close to the patient and decision aids that are easily understood by considering people’s HL and graphic literacy with acceptable words, phrases, and images to complement functional HL. [14] This is desired to understand the multi-item tests and their symptoms [12] and to take advantage of the Japanese system of application for intractable diseases to exempt patients from medical expenses. As shown in the present study, rheumatologists may need to be attentive to whether functional HL should be augmented in older or depressed individuals.

Second, communicative HL may be facilitated by training rheumatologists in dialogue approaches to promote patients’ self-esteem and efficacy. Preventing discrepancies between what the rheumatologist recommends and what the patient understands, for example, the significance of tapering steroids while taking them correctly, can be confirmed by teach-back and activating such dialogue can improve communicative HL. For this purpose, an atmosphere where patients can talk to their rheumatologist about what they understand and an environment where rheumatologists show interest in their disease without hesitation, whether it is appropriate or not, needs to be created. The use of a toolkit for HL reminders with encouraging questions and teach-backs for patients with rheumatoid arthritis has been proven to increase adherence to treatment. [15]

Our study has several strengths. First, this is the first study to reveal an association between trust in one’s own physician and HL among patients with SLE. In addition, we were able to expand the correlates of HL by measuring broader concepts of HL dimensions, such as communicative and critical HL. It is also worth mentioning that we were able to show the association between HL and trust in one’s physician by studying a single race, eliminating racial differences, and adjusting for educational and economic status. Second, the multicenter design ensured the external validity of our findings.

Although this study demonstrated vital insights, it has several limitations. First, HL was not measured using an objective test, although a self-report instrument with well-validated reliability and validity was used. However, Japan, like the U.S., has a literacy rate of 99% [16]. Second, the duration of Internet use was not limited to the search for health information. However, the duration of Internet use may reflect the ability to access online health information. Third, due to the cross-sectional nature of this study, attention should be paid to reverse causality. An alternative explanation could be that the loss of trust in physicians increases the motivation to acquire health information on one’s own or generates a belief in interpreting health information carefully.

In conclusion, in patients with SLE, higher trust in one’s physician was associated with both functional and communicative HL, while higher trust in physicians in general was associated with higher communicative and lower critical HL. Our findings suggest that trusting relationships may be fostered if rheumatologists encourage patients to share their health problems with their physicians and family members and to obtain useful health information (i.e., communicative HL), rather than focusing on improving their ability to discern health information (i.e., critical HL).

## Data Availability

The datasets generated during and/or analyzed during the study are available from the corresponding author on reasonable request.

## ACKNOWLEDGMENTS

We would especially like to thank Prof Hirono Ishikawa for her advice and support with the FCCHL scale, Hiroko Nagasato, Kumi Sasaki, Yukari Hosaka (Showa University), and Miyuki Sato (Fukushima Medical University) for their clerical support. Part of this study was presented at the 66th Annual General Assembly and Scientific Meeting of the Japan College of Rheumatology.

