## Supplementary Table 1 for "The impact of communicative and critical health literacy on trust in physicians among patients with systemic lupus erythematosus: the TRUMP2-SLE project"

**Supplementary Table 1 The Functional Communicative Critical Health Literacy Scale (FCCHL)**

| | |
|---|---|
| Instruction sentence | あなたは、この1年間に、病院や薬局からもらう説明書やパンフレットを読む際、次のようなことがありましたか。最も当てはまる番号に○をしてください。<br>(English: “In reading instructions or leaflets from hospitals/pharmacies, you. . .”) |
| Question 1 | 字が細かくて、読みにくい（メガネなどをかけた状態でも）<br>(English: “found that the print was too small to read.”) |
| Question 2 | 読めない漢字や知らない言葉がある。<br>(English: “found characters and words that you did not know”) |
| Question 3 | 内容が難しくて分かりにくい<br>(English: “found that the content was too difficult.”) |
| Question 4 | 読むのに時間がかかる<br>(English: “needed a long time to read and understand them.”) |
| Question 5 | 誰かに代わりに読んで教えてもらう<br>(English: “needed someone to help you read them.”) |
| Instruction sentence | 全身性エリテマトーデスと診断されてから、全身性エリテマトーデスやその治療・健康法に関することについて、以下のようなこと |

| | |
|---|---|
| | をしましたか。最も当てはまる番号に○をしてください。<br><br>(English: “Since having systemic lupus erythematosus you have. . .”) |
| Question 1 | いろいろなところから知識や情報を集めた<br><br>(English: “collected information from various sources.”) |
| Question 2 | たくさんある知識や情報から、自分の求めるものを選び出した<br><br>(English: “extracted the information you wanted.”) |
| Question 3 | 自分が見聞きした知識や情報を、理解できた<br><br>(English: “understood the obtained information.”) |
| Question 4 | 病気についての自分の意見や考えを、医師や身近な人に伝えた<br><br>(English: “communicated your thoughts about your illness to someone.”) |
| Question 5 | 見聞きした知識や情報をもとに、実際に生活を変えてみた<br><br>(English: “applied the obtained information to your daily life.”) |
| Question 6 | 見聞きした知識や情報が、自分にもあてはまるかどうか考えた<br><br>(English: “considered whether the information was applicable to your situation.”) |
| Question 7 | 見聞きした知識や情報の信頼性に疑問をもった<br><br>(English: “considered the credibility of the information.”) |

| | |
|---|---|
| Question 8 | 見聞きした知識や情報が正しいかどうか聞いたり、調べたりした<br><br>(English: "checked whether the information was valid and reliable.") |
| Question 9 | 病気や治療法などを自分で決めるために調べた<br><br>(English: "collected information to make health-related decisions.") |
| Response options for questions | 全くなかった/あまりなかった/時々あった/よくあった<br><br>(English: "never / rarely / sometimes / often") |

The English-translated version [1] is also provided for each item and response.
